# Childhood-onset of primary Sjögren syndrome Phenotypic characterization at diagnosis of 158 children

**DOI:** 10.1101/2020.05.12.20099168

**Authors:** Manuel Ramos-Casals, Nihan Acar-Denizli, Arjan Vissink, Pilar Brito-Zerón, Xiaomei Li, Francesco Carubbi, Roberta Priori, Nataša Toplak, Chiara Baldini, Enrique Faugier-Fuentes, Aike A. Kruize, Thomas Mandl, Minako Tomiita, Saviana Gandolfo, Kunio Hashimoto, Gabriela Hernandez-Molina, Benedikt Hofauer, Samara Mendieta-Zerón, Astrid Rasmussen, Pulukool Sandhya, Damien Sene, Virginia Fernandes Moça Trevisani, David Isenberg, Erik Sundberg, Sandra G. Pasoto, Agata Sebastian, Yasunori Suzuki, Soledad Retamozo, Bei Xu, Roberto Giacomelli, Angelica Gattamelata, Masa Bizjak, Stefano Bombardieri, Richard-Eduardo Loor-Chavez, Eefje van der Heijden, Peter Olsson, Hendrika Bootsma, Scott M. Lieberman, for the Sjogren Big Data Consortium

**Affiliations:** Department of Autoimmune Diseases, ICMiD, Hospital Clínic, Barcelona, Spain; Sjögren Syndrome Research Group (AGAUR), Laboratory of Autoimmune Diseases Josep Font, IDIBAPS- CELLEX, Barcelona, Spain; Department of Medicine, University of Barcelona, Barcelona, Spain; Department of Statistics, Faculty of Science and Letters, Mimar Sinan Fine Arts University, Istanbul, Turkey; Department of Oral and Maxillofacial Surgery, University of Groningen, University Medical Center Groningen, Groningen, The Netherlands; Autoimmune Diseases Unit, Department of Medicine, Hospital CIMA-Sanitas, Barcelona, Spain; Department of Rheumatology and Immunology, Anhui Provincial Hospital, Hefei, China; Clinical Unit of Rheumatology, University of I’Aquila, School of Medicine, L’Aquila, Italy; Department of Internal Medicine and Medical Specialties, Rheumatology Clinic, Sapienza University of Rome, Rome, Italy; University Children’s Hospital Ljubljana, University Medical center Ljubljana, Medical faculty of Ljubljana, Slovenia; Rheumatology Unit, University of Pisa, Pisa, Italy; Hospital Infantil de México Federico Gómez, Ciudad de México, México; Department of Rheumatology and Clinical Immunology, University Medical Center Utrecht, Utrecht, The Netherlands; Department of Rheumatology, Skane University Hospital Malmö, Lund University, Lund, Sweden; Department of Pediatrics, National Hospital Organization, Shimoshizu National Hospital, 943-5 Shikawatashi, Yotsukaido, Chiba, Japan; Clinic of Rheumatology, Department of Medical and Biological Sciences, University Hospital “Santa Maria della Misericordia”, Udine, Italy; Nagasaki University Graduate School of Biomedical Sciences, Department of Pediatrics (Pediatric Allergy and Rheumatology), Nagasaki, Japan; Immunology and Rheumatology Department, Instituto Nacional de Ciencias Médicas y Nutrición Salvador Zubirán. México City, Mexico; Otorhinolaryngology, Head and Neck Surgery, Technical University Munich, Munich, Germany; Hospital Materno Infantil ISSEMyM Toluca, México; Arthritis and Clinical Immunology Research Program, Oklahoma Medical Research Foundation, Oklahoma City, OK, USA; Department of Clinical Immunology & Rheumatology, Christian Medical College & Hospital, Vellore, India; Service de Médecine Interne 2, Hôpital Lariboisière, Université Paris VII, Assistance Publique-Hôpitaux de Paris, 2, Paris, France; Federal University of São Paulo, Sao Paulo, Brazil; Centre for Rheumatology, Division of Medicine, University College London, London, UK; Pediatric Rheumatology, Astrid Lindgren’s Children Hospital, Karolinska Institutet, and Karolinska University Hospital, Stockholm, Sweden; Rheumatology Division, Hospital das Clinicas, Faculdade de Medicina da Universidade de Sao Paulo (HCFMUSP), Sao Paulo, Brazil; Department of Rheumatology and Internal Medicine, Wroclaw Medical University, Wroclaw, Poland; Division of Rheumatology, Kanazawa University Hospital, Kanazawa, Ishikawa, Japan; Instituto De Investigaciones En Ciencias De La Salud (INICSA), Universidad Nacional de Córdoba (UNC), Consejo Nacional de Investigaciones Científicas y Técnicas (CONICET) - Córdoba - Argentina. Instituto Universitario de Ciencias Biomédicas de Córdoba (IUCBC), Córdoba- Argentina; Department of Rheumatology and Clinical Immunology, University of Groningen, University Medical Center Groningen, Groningen, The Netherlands; Stead Family Department of Pediatrics, Carver College of Medicine, University of Iowa, Iowa City, Iowa, USA

**Keywords:** Sjogren’s Syndrome, Epidemiology, Autoimmune Diseases, Paediatrics, Childhood

## Abstract

**OBJECTIVES:** To characterize the phenotypic presentation at diagnosis of childhood-onset primary Sjögren syndrome (SjS).

**METHODS:** The Big Data Sjögren Project Consortium is an International, multicentre registry using worldwide data-sharing cooperative merging of pre-existing clinical SjS databases from the five continents. For this study, we selected those patients in whom the disease was diagnosed below the age of 19 according to the fulfilment of the 2002/2016 classification criteria.

**RESULTS:** Among the 12,083 patients included in the Sjögren Big Data Registry, 158 (1.3%) patients had a childhood-onset diagnosis (136 girls, mean age of 14.2 years): 126 (80%) reported dry mouth, 111 (70%) dry eyes, 52 (33%) parotid enlargement, 118/122 (97%) positive minor salivary gland biopsy and 60/64 (94%) abnormal salivary US study, 140/155 (90%) positive ANA, 138/156 (89%) anti-Ro/La antibodies and 86/142 (68%) positive RF. The systemic ESSDAI domains containing the highest frequencies of active patients included the glandular (47%), articular (26%) and lymphadenopathy (25%) domains. Patients with childhood-onset primary SjS showed the highest mean ESSDAI score and the highest frequencies of systemic disease in 5 (constitutional, lymphadenopathy, glandular, cutaneous and haematological) of the 12 ESSDAI domains, and the lowest frequencies in 4 (articular, pulmonary, peripheral nerve and central nervous system) in comparison with patients with adult-onset disease.

**CONCLUSIONS:** Childhood-onset primary SjS involves around 1% of patients with primary SjS, with a clinical phenotype dominated by sicca features, parotid enlargement and systemic disease. Age at diagnosis plays a key role on modulating the phenotypic expression of the disease.

## INTRODUCTION

Primary Sjögren syndrome (SjS) is a systemic autoimmune disease most commonly diagnosed in middle-aged women, with a frequency ranging between 0.01 and 0.72% [1], The autoimmune damage mainly targets the exocrine glands, which are infiltrated by lymphocytes (focal sialadenitis) [2], More than 95% of patients present with oral and/or ocular dryness [3], but may also develop a large number of organ-specific manifestations (systemic Sjögren) [4], The key immunological markers are anti-Ro antibodies, the most specific, and cryoglobulins and hypocomplementaemia, the main prognostic markers [5].

Although primary SjS can occur at all ages, it is diagnosed between 30–60 years in two thirds of patients [3]. Paediatric onset of the disease is rarely reported, and there is no information about how frequent the paediatric presentation is. In addition, a clear view of how the disease presents in children is difficult to obtain due to the scarce and heterogeneous available data. The main series published until now are characterized by a heterogeneous methodology, using a variable definition of the paediatric age (from 14 to 18 years), including up to 50% of patients not fulfilling the current classification criteria and mixing the inclusion of primary and associated forms of the disease. As an example, in the largest series reported (67 cases) [6], only approximately half the cases fulfilled the current SjS classification criteria and 12% were patients diagnosed with other concomitant systemic autoimmune diseases.

Understanding how primary SjS presents/manifests at earlier ages (both glandular and extraglandular) would help Paediatricians to more promptly identify the disease [7], In the absence of established classification criteria specific for SjS in children, along with the notions that SjS in children is not pathophysiologically distinct from SjS in adults and that childhood-onset primary SjS is part of the epidemiological continuum of the disease, we chose to focus our study on children who met the current classification criteria [8,9] established for use in classifying SjS in adults.

Therefore, the aim of this study was to characterize the phenotypic presentation at diagnosis of childhood-onset primary SjS in a large international, multi-ethnic cohort of patients in comparison with the adult-onset phenotype.

## METHODS

### Patients

The Big Data Sjögren Project Consortium is an international, multicentre registry designed in 2014 to take a “high-definition” picture of the main features of primary SjS using worldwide data-sharing cooperative merging of pre-existing clinical SjS databases from leading centres in clinical research in SjS from the five continents [3,10,11]. Databases from each centre are harmonized into a single data base by applying the data-cleaning pre-processing techniques. Descriptive statistics and data visualisation methods are used in order to detect outliers, data errors, missing data and influential observations [12]. A double-checking process correcting errors and completing missing information is carried out to minimize incomplete and erroneous data. Inclusion criteria are the fulfilment of the 2002 classification criteria [8] and/or 2016 ACR/EULAR criteria [9]. Diagnostic tests for SjS were carried out according to the recommendations of the European Community Study Group [13]. The study was approved by the Ethics Committee of the Coordinating Centre (Hospital Clinic, Barcelona, Spain, registry HCB/2015/0869).

### Definition of variables

Disease diagnosis was defined as the time when the attending physician confirmed fulfilment of the 2002/2016 criteria. For this study, we selected those patients in whom the disease was diagnosed below the age of 19. Because the members of the Project are physicians that care for adult patients, the coordinator of each center travelled to the referral paediatric centers to include paediatric cases with active follow-up, in order to avoid selection bias by excluding younger children who were not yet referred to the adult departments.

The main disease features at this time (criteria fulfilment), as well as the first signs and symptoms at presentation, were retrospectively collected from the patient records and analysed. The following clinical variables were selected for harmonization and further refinement:

a. Epidemiological features: age, gender, ethnicity, country of residence.
b. Clinical features at presentation: first signs and symptoms suggesting an autoimmune disease.
c. Systemic activity: systemic involvement at diagnosis was classified and scored according to the EULAR Sjögren’s syndrome disease activity index (ESSDAI) [14], the clinical ESSDAI (clinESSDAI) [15] [16] and the disease activity states (DAS) [17].
d. Diagnostic approach: fulfilment of the 2002/2016 criteria items, parotid ultrasound study.
e. Immunological profile: antinuclear antibodies, anti-Ro/La antibodies, rheumatoid factor, complement C3 and C4 levels, cryoglobulins.

### Statistical analysis

Descriptive data are presented as mean and standard deviation (SD) for continuous variables and numbers and percentages (%) for categorical variables. Disease patterns for three paediatric age at diagnosis subsets (5-9 years, 10-14 years, 15-18 years) and five 18-year age at diagnosis groups (≤18 years, 19-36 years, 37-54 years, 55-72 years, ≥73 years) were compared according to the gender, ethnicity, diagnostic tests for SjS, immunological markers and systemic involvement. The Chi-square test was used to compare categorical variables and ANOVA test was used to compare continuous variables. Data visualization techniques were used to summarize information. To handle missing data due to non-evaluated features, “available case analysis” was assumed. All significance tests were two-tailed and values of p < 0.05 were considered significant. P-values were adjusted for multiple comparisons using the false discovery rate (FDR) correction. All analyses were conducted using the R V.3.5.0 for Windows statistical software package (https://www.R-project.org/).

## RESULTS

Among the 12,083 patients included in the Sjögren Big Data Registry, 158 (1.3%) patients were diagnosed at an age below 19 years **(Figure 1)**. They were 136 (86%) girls and 22 (14%) boys, with a mean age at first sign or symptom suggestive of the disease of 13.2 (SD 3.2) years and of 14.2 (SD 3.5) years at the time of diagnosis of primary SjS **(Table 1)**. The first signs and symptoms that led to the suspicion of an autoimmune disease are summarized in **Table 1**.

**Figure 1.**
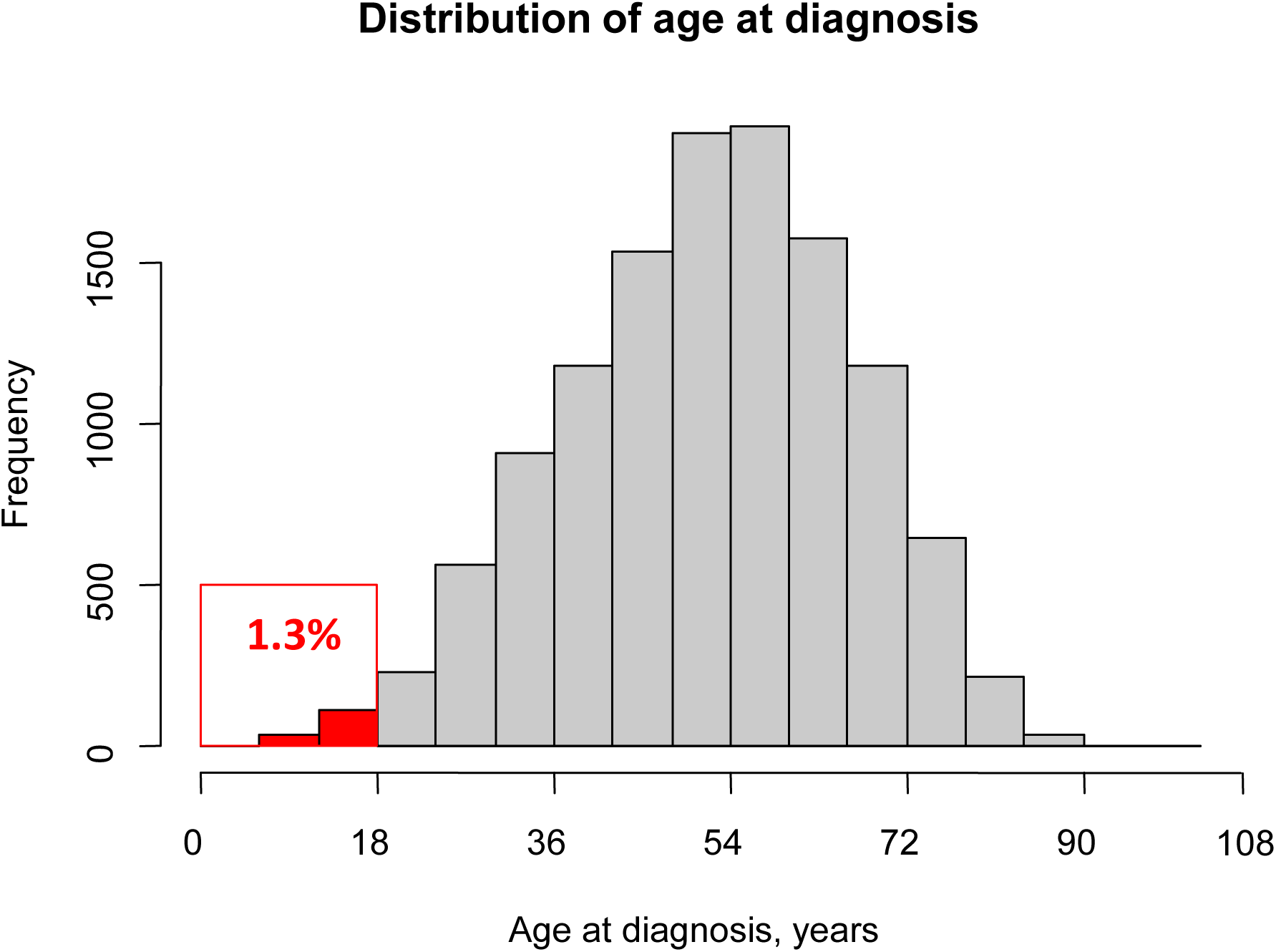
Histogram representing the distribution of age at diagnosis in five 18-year age groups (≤18 years, 19-36 years, 37-54 years, 55-72 years, ≥73 years).

**Table 1.** Epidemiological and clinical features of 158 patients with primary Sjögren syndrome diagnosed in childhood.

| **Variable** | **Patients with pSjS**<br>**diagnosed in**<br>**childhood (n = 158)** |
| --- | --- |
| Sex and age |  |
| Sex (female) | 136 (86.1) |
| Age at diagnosis (mean ± SD) | 14.2 ± 3.5 |
| Age at first sign/symptom (n = 29) | 13.2 ± 3.2 |
| Ethnicity |  |
| White | 107 (67.7) |
| Asian | 26 (16.5) |
| Hispanic | 19 (12) |
| Black/African-American (AA) | 4 (2.5) |
| Others | 2 (1.3) |
| Geolocation |  |
| Europe | 104 (65.8) |
| America | 29 (18.4) |
| Asia | 24 (15.2) |
| Africa | 1 (0.6) |
| Signs & symptoms at presentation |  |
| Glandular enlargement | 54/151 (35.8) |
| Dry mouth + dry eyes | 36/151 (23.8) |
| Isolated dry mouth or dry eyes | 33/151 (21.9) |
| Fever | 18/151 (11.9) |
| Arthralgias | 15/151 (9.9) |
| Skin involvement | 14/151 (9.3) |
| Fatigue | 7/151 (4.6) |
| Raynaud phenomenon | 7/151 (4.6) |
| Peripheral lymphadenopathies | 5/151 (3.3) |
| Arthritis | 4/151 (2.6) |
| Cytopenias | 4/151 (2.6) |
| RTA | 2/151 (1.3) |
| Myalgias | 1/151 (0.7) |

Despite not being presenting complaints, 126 (80%) children reported dry mouth and 111 (70%) dry eyes at the time of diagnosis. With respect to the diagnostic approach, abnormal ocular tests were reported in 51% in those studied by Schirmer’s test and 35% in those studied using ocular dye tests, while abnormal oral tests were reported in 78% of those studied by unstimulated whole salivary flows (UWSF) and 81% of those studied by parotid scintigraphy **(Table 2)**. Minor salivary gland biopsy showed Chisholm Mason grades 3-4 (ie, focus score ≥ 1 focus/4 mm^2^) in 118 (97%) out of 122 patients biopsied, while salivary US study was abnormal in 60 (94%) out of 64 patients **(Table 2)**. With respect to the immunological profile at the time of diagnosis, the most frequent abnormalities consisted of positive ANA (90%), anti-Ro antibodies (83%), positive rheumatoid factor (RF) (68%) and anti-La antibodies (62%) **(Table 2)**. All patients fulfilled both the 2002 and 2016 criteria except 3 (2%) who had anti-La antibodies in the absence of anti-Ro antibodies and in whom fulfilment of the 2016 criteria could not be confirmed because a salivary gland biopsy was not performed.

With respect to systemic phenotype, the mean total ESSDAI score at diagnosis of the entire cohort was 7.1 (SD 6.7); 90% of patients had systemic activity at diagnosis (global ESSDAI score ≥ 1) **(Table 2)**. The clinical domains containing the highest frequencies of active patients included the glandular (47%), articular (26%) and lymphadenopathy (25%) domains. There was only 1 (0.6%) patient who presented with lymphoma at the time of primary SjS diagnosis. The distribution of the degree of activity (no activity, low, moderate and high) in the entire cohort for each domain is summarized in **Supplementary Table 1**, and the association of the main variables at diagnosis with systemic activity is summarized in **Supplementary Table 2**.

**Table 2.** Diagnostic tests, immunological markers and systemic activity at the time of diagnosis of 158 patients with primary Sjögren syndrome diagnosed in childhood.

| Variable | Patients with pSjS<br>diagnosed in childhood (n = 158) |
| --- | --- |
| **DIAGNOSTIC TESTS** |  |
| Abnormal ocular tests (any) | 72/125 (57.6) |
| Schirmer's test | 63/123 (51.2) |
| Rose bengal score/other ocular dye score | 27/76 (35.5) |
| Abnormal oral diagnostic tests (any) | 80/98 (81.6) |
| Unstimulated whole salivary flow | 62/79 (78.5) |
| Parotid sialography | 16/18 (88.9) |
| Salivary scintigraphy | 17/21 (81) |
| Positive minor salivary gland biopsy | 118/122 (96.7) |
| Abnormal salivary US study | 60/64 (93.8) |
| **IMMUNOLOGICAL PROFILE** |  |
| ANA-positive | 140/155 (90.3) |
| RF-positive | 96/142 (67.6) |
| Positive anti-Ro/La antibodies | 138/156 (88.5) |
| Anti-Ro antibodies | 129/156 (82.7) |
| Anti-La antibodies | 96/155 (61.9) |
| Low C3 levels | 20/138 (14.5) |
| Low C4 levels | 19/136 (14) |
| Positive cryoglobulins | 3/64 (4.7) |
| **SYSTEMIC ACTIVITY** |  |
| Mean ESSDAI score (n=155) | 7.1 ± 6.7 |
| DAS |  |
| Low | 72/155 (46.5) |
| Moderate | 57/155 (36.8) |
| High | 26/155 (16.8) |
| Activity subsets |  |
| No activity (ESSDAI = 0) | 16/155 (10.3) |
| No high activity in any domain | 128/155 (82.6) |
| High activity in at least 1 domain | 11/155 (7.1) |
| ESSDAI domains (score ≥1) |  |
| Constitutional | 34/155 (21.9) |
| Lymphadenopathy | 39/155 (25.2) |
| Glandular | 73/155 (47.1) |
| Articular | 41/155 (26.5) |
| Cutaneous | 19/155 (12.3) |
| Pulmonary | 8/155 (5.2) |
| Renal | 7/155 (4.5) |
| Muscular | 3/155 (1.9) |
| PNS | 0/155 (0) |
| CNS | 1/155 (0.6) |
| Haematological | 44/155 (28.4) |
| Biological | 84/155 (54.2) |

**Table 3** compares the disease patterns at diagnosis according to the 3 paediatric age subsets (<10 years, 10-14 years, >14 years). These analyses revealed some similarities and some differences including the increase in Whites (p=0.003), a higher frequency of reported dry eyes (p=0.026) and a lower frequency in lymphadenopathy (p=0.003) in the older group compared to the other two younger groups. In addition, patients diagnosed at 10-14 years showed the highest systemic activity phenotype, with a higher frequency of anti-Ro antibodies (p=0.042), a higher global mean ESSDAI (p=0.017) and a higher frequency of systemic activity at the constitutional domain (p=0.004). Using 15 as the cutoff resulted in increased frequency of low C3 levels (p=0.049) and an increased frequency of patients with no systemic activity (p=0.012) in the older compared to younger group **(Supplementary Table 3)**.

**Table 3.**
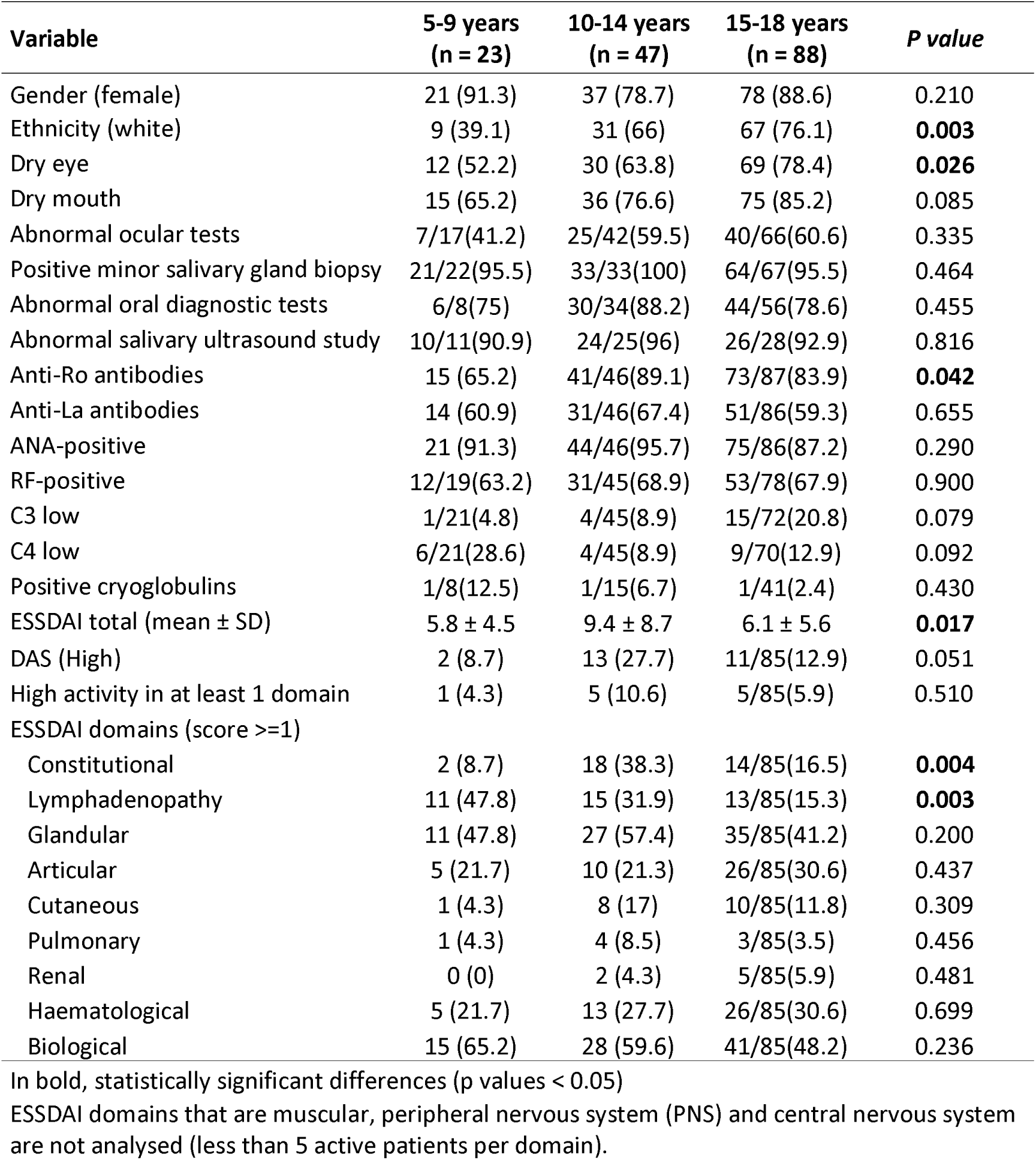
Differences in the pattern expression of primary SjS according to the three paediatric age subsets (5-9 years, 10-14 years and 15-18 years).

**Table 4** compares the disease pattern of childhood-onset primary SjS with adult patients grouped in intervals of 18 years. Childhood-onset primary SjS showed a higher frequency of affected male sex and non-White patients, a lower frequency of sicca features with a lower frequency of abnormal ocular tests but a higher frequency of abnormal oral tests and positive salivary gland biopsy **(Figure 2A)**. With respect to immunological profile, childhood-onset disease patients showed the highest frequencies of positive autoantibodies and the lowest frequency of cryoglobulins among all the age subsets **(Figure 2B)**. With respect to systemic disease, the highest mean ESSDAI score was observed in childhood patients, who also showed the highest frequencies of systemic disease in 5 (constitutional, lymphadenopathy, glandular, cutaneous and haematological) out of the 12 ESSDAI domains, and the lowest frequencies in 4 (articular, pulmonary, peripheral nerve and central nervous system) **(Figure 2C)**. There is a differentiated trend in the frequency of patients with active organ-specific ESSDAI domains according to the age group at diagnosis; while some organs follow a predominant steady decreasing frequency pattern (constitutional, lymphadenopathy and glandular), others showed a predominant steady increasing pattern (pulmonary and neuromuscular) from the youngest to the older group ages **(Figure 3)**.

**Figure 2.**
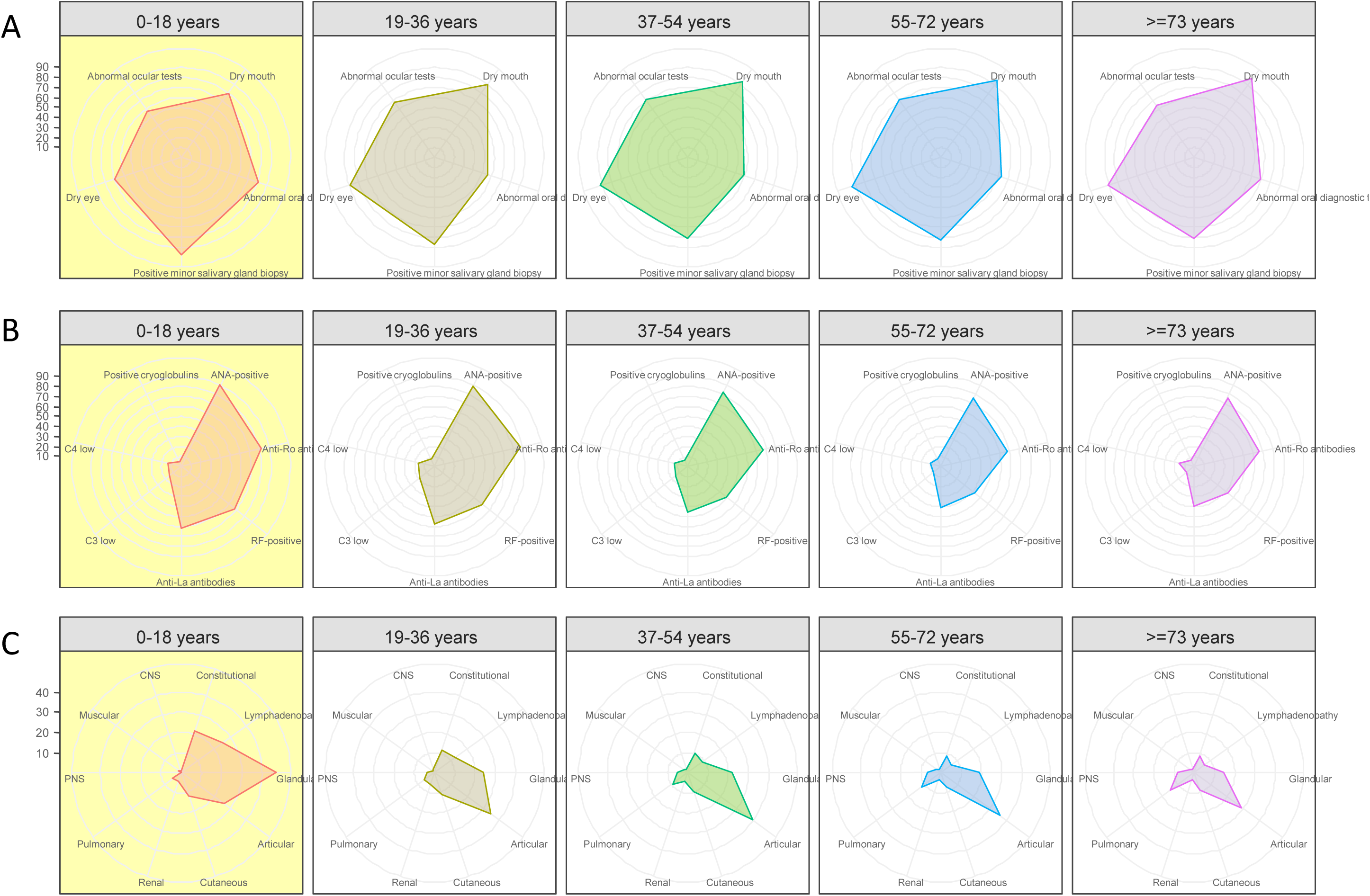
Radar charts showing variations in glandular involvement (A), immunological profile (B) and systemic disease using the ESSDAI classification (C) between age at diagnosis groups (≤18 years, 19-36 years, 37-54 years, 55-72 years, ≥73 years).

**Figure 3.**
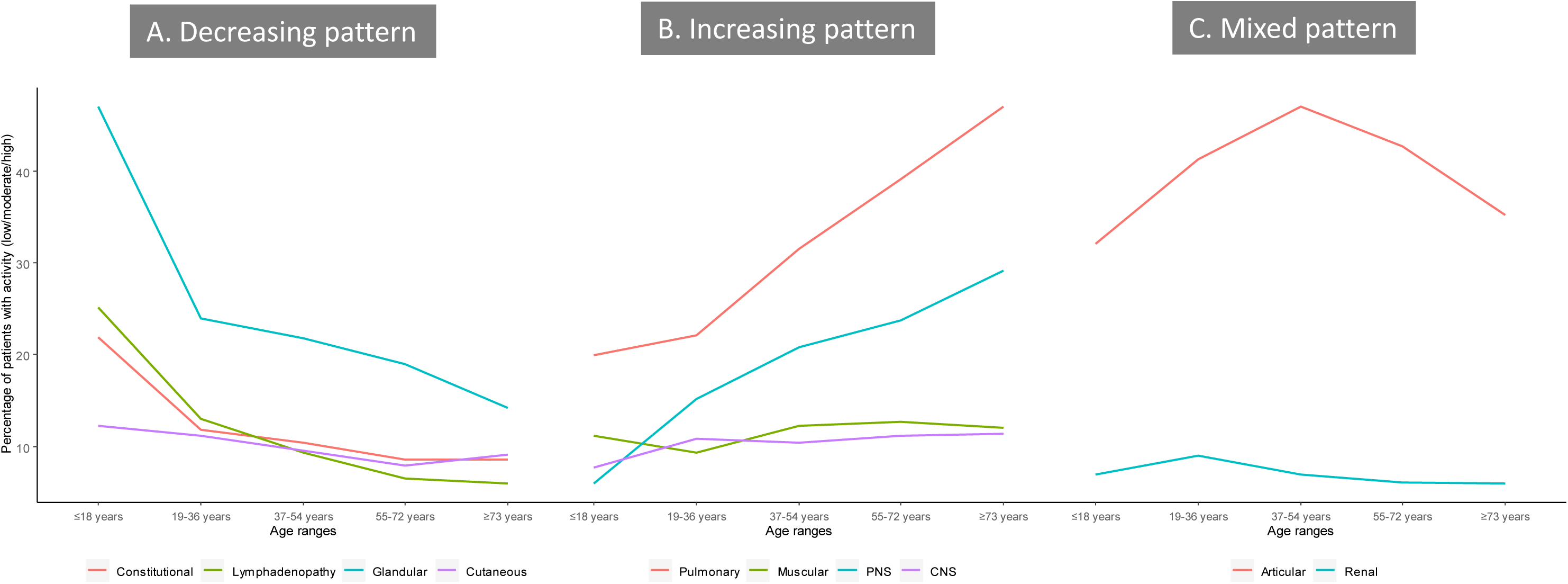
Frequency of patients with active organ-specific ESSDAI domains according to the five 18-year age groups (≤18 years, 19-36 years, 37-54 years, 55-72 years, ≥73 years). (A) Organs with a predominant decreasing pattern; (B) Organs with a predominant increasing pattern; (C): Organs with a mixed pattern.

**Table 4.** Differences in the pattern expression of primary SjS according to the five 18-year age groups (≤18 years, 19-36 years, 37-54 years, 55-72 years, ≥73 years).

| Variable | $\leq 18$ years<br>(n = 158) | 19-36 years<br>(n = 1701) | 37-54 years<br>(n = 4591) | 55-72 years<br>(n = 4684) | $\geq 73$ years<br>(n = 921) |
| --- | --- | --- | --- | --- | --- |
| Gender (female) | 136 (86.1) | 1625 (95.5) | 4302 (93.7) | 4361 (93.1) | 845 (91.7) |
| Ethnicity (white) | 107 (67.7) | 1073/1674 (64.1) | 3130/4476 (69.9) | 3529/4468 (79) | 756/877 (86.2) |
| Dry eye | 111 (70.3) | 1498 (88.1) | 4229 (92.1) | 4403 (94) | 840 (91.2) |
| Dry mouth | 126 (79.7) | 1532 (90.1) | 4282 (93.3) | 4447 (94.9) | 892 (96.9) |
| Abnormal ocular tests | 72/125 (57.6) | 1151 (67.7) | 3298 (71.8) | 3342 (71.3) | 592 (64.3) |
| Positive minor salivary gland biopsy | 118/122 (96.7) | 1001/1160 (86.3) | 2601/3223 (80.7) | 2868/3501 (81.9) | 537/663 (81) |
| Abnormal oral diagnostic tests | 80/98 (81.6) | 958 (56.3) | 2681 (58.4) | 2962 (63.2) | 637 (69.2) |
| Anti-Ro antibodies | 129/156 (82.7) | 1480/1682 (88) | 3505/4535 (77.3) | 3172/4641 (68.3) | 604/919 (65.7) |
| Anti-La antibodies | 96/155 (61.9) | 953/1674 (56.9) | 2072/4503 (46) | 1874/4626 (40.5) | 359/915 (39.2) |
| ANA-positive | 140/155 (90.3) | 1472/1640 (89.8) | 3594/4356 (82.5) | 3289/4326 (76) | 645/845 (76.3) |
| RF-positive | 96/142 (67.6) | 820/1348 (60.8) | 1846/3752 (49.2) | 1580/3665 (43.1) | 309/725 (42.6) |
| C3 low | 20/138 (14.5) | 259/1406 (18.4) | 563/3766 (14.9) | 349/3715 (9.4) | 73/727 (10) |
| C4 low | 19/136 (14) | 232/1398 (16.6) | 535/3763 (14.2) | 435/3712 (11.7) | 109/733 (14.9) |
| Positive cryoglobulins | 3/64 (4.7) | 60/730 (8.2) | 134/2055 (6.5) | 177/2068 (8.6) | 28/388 (7.2) |
| ESSDAI total (mean ± SD) | 7.1 ± 6.7 | 6.2 ± 6.6 | 6.0 ± 7.3 | 5.8 ± 7.7 | 6.1 ± 7.4 |
| ESSDAI domains (score $\geq 1$) | | | | | |
| Constitutional | 34/155(21.9) | 191/1621(11.8) | 458/4416(10.4) | 380/4430(8.6) | 74/860(8.6) |
| Lymphadenopathy | 39/155(25.2) | 211/1625(13) | 410/4423(9.3) | 290/4440(6.5) | 52/863(6) |
| Glandular | 73/155(47.1) | 389/1621(24) | 961/4416(21.8) | 846/4430(19.1) | 121/860(14.1) |
| Articular | 41/155(26.5) | 558/1621(34.4) | 1743/4416(39.5) | 1585/4430(35.8) | 250/859(29.1) |
| Cutaneous | 19/155(12.3) | 181/1625(11.1) | 422/4423(9.5) | 349/4440(7.9) | 78/863(9) |
| Pulmonary | 8/155(5.2) | 97/1625(6) | 420/4422(9.5) | 545/4440(12.3) | 130/863(15.1) |
| Renal | 7/155(4.5) | 101/1621(6.2) | 196/4416(4.4) | 162/4430(3.7) | 32/860(3.7) |
| Muscular | 3/155(1.9) | 20/1621(1.2) | 102/4416(2.3) | 111/4430(2.5) | 19/860(2.2) |
| PNS | 0/155(0) | 55/1621(3.4) | 240/4416(5.4) | 293/4430(6.6) | 74/860(8.6) |
| CNS | 1/155(0.6) | 29/1621(1.8) | 72/4416(1.6) | 84/4430(1.9) | 17/860(2) |
| Haematological | 44/155(28.4) | 387/1611(24) | 981/4365(22.5) | 858/4368(19.6) | 200/843(23.7) |
| Biological | 84/155(54.2) | 1009/1584(63.7) | 2205/4276(51.6) | 1911/4301(44.4) | 370/846(43.7) |

All comparisons were statistically significant (p-values $< 0.05$ ) except for positive cryoglobulins (p=0.117), total ESSDAI (p=0.069), muscular domain (p=0.058) and CNS (p=0.691).

## DISCUSSION

A significant challenge of diagnosing primary SjS is recognizing the disease can occur in people of any age. Although the disease is diagnosed predominantly in middle-aged people, the epidemiology of primary SjS is a continuum and the disease has been diagnosed in people from 2 [18] to 97 years [10]. The data available until now suggest that primary SjS is a very rare disease in children. In comparison with adult-onset disease, both the number of studies and the number of reported cases of childhood-onset SjS is very limited, with only 6 studies including more than 20 cases [6,19-23] **(Supplementary Table 4)**. In this study, we use a different methodological approach from previous studies. Firstly, we included only cases fulfilling the current classification criteria for the disease [8,9] searching for those cases included in the Sjögren Big Data cohort diagnosed in childhood. And secondly, we requested the cooperative involvement of paediatricians who included children currently followed in paediatric departments for ensuring a complete epidemiological coverage of potential cases. For the first time, we can estimate the real frequency of childhood-onset disease (around 1%), confirming that primary SjS is a disease that is exceptionally diagnosed in children. Previous studies have defined the clinical phenotype of childhood-onset SS as mainly dominated by parotid involvement more than by sicca features, often using a clinical diagnosis more than the current SjS classification criteria **(Supplementary Table 4)**. Our study, made under the methodological umbrella of the fulfilment of current criteria, showed a predominant phenotype much closer to that reported in young adult patients, with sicca symptoms being more frequent than salivary gland enlargement (80% and nearly 50%, respectively). Despite this, salivary gland enlargement remains a prominent feature of childhood-onset primary SjS affecting one out of two children. In these children, it is important to distinguish onset of SjS from recurrent parotitis of childhood (RP), an idiopathic disease affecting the parotid glands reported as the most common parotid disease in childhood after mumps [24], There are significant differences between RP and SjS, especially in the immunological profile (ANA, RF, Ro and/or La antibodies) that was reported in 96% of our cases of childhood primary SjS, while in RP patients, the frequency ranges from 4% to 16% [25,26].

Classification criteria for SS in children are not uniformly agreed upon by paediatricians [27] and diagnostic tests used for diagnosing glandular dysfunction in adults are not validated in children. Among ocular tests, Schirmer’s test is most frequently carried out in children with SjS, with around 70% of cases showing abnormal results [19,23]. For the study of salivary gland dysfunction, salivary gland ultrasound (SGUS) is a non-invasive test that may evaluate the glandular damage of salivary glands and that can be very useful in both adults and children. In the largest study reported, Hammenfors et al [6] have evaluated 67 patients with childhood-onset SjS in whom ultrasound was carried out at a mean age of 16 years (including an undetermined number of cases evaluated above the age of 18): pathologic SGUS findings were observed in 41 (61%) cases, while in our study, the rate was higher (94%). The low rate of positive findings reported by Hammenfors et al [6] could be explained by the fact that less than 60% of cases fulfilled the current classification criteria, as well as by the significant variation among countries. But the two key diagnostic markers supporting a diagnosis of SjS are, as happens in adults, a positive test for Ro autoantibodies and the finding of a focal lymphocytic sialadenitis in the salivary gland tissue, because they are the most specific diagnostic tests. In previous studies, salivary gland biopsy was positive in 82% of children **(Supplementary Table 4)**, while in our study the frequency raised to 97%. With respect to immunological markers, anti-Ro/SSA antibodies were found in more than 70% of patients and anti-Ro/SSA and/or anti-La/SSB in 80% of childhood SS patients in previous studies [27], In our study, nearly 90% of our cases were positive for Ro/La antibodies, highlighting the key role of these autoantibodies in diagnosing primary SjS in children.

According to our data, childhood primary SjS is undeniably a systemic disease (90% of our cases had systemic activity at the time of diagnosis, defined as an ESSDAI score of 1 or higher), as happens in adults. The age at diagnosis is a key determinant of the expression of systemic disease in primary SjS, and a recent study in our international cohort showed that the highest systemic scores were reported in patients diagnosed at 18-35 years [10]. In addition, age at diagnosis also modulated how frequent the involvement of individual organs was at the time of diagnosis. A younger diagnosis was associated with an increased risk of presenting activity at diagnosis in some clinical domains (constitutional, lymphadenopathy, glandular, cutaneous, renal), but a decreased risk of presenting activity in others (pulmonary and neurological domains). In childhood-onset primary SjS, systemic phenotype is clearly dominated by glandular involvement (in nearly 50% of our patients), followed by articular, lymphadenopathy and constitutional involvement, a very close phenotypic scenario to that reported in young-onset people [10].

The inclusion criteria that we used in this study (primary cases meeting the current criteria) may explain some of the main findings, such as the higher percentages of positive diagnostic tests (salivary flow, salivary gland ultrasound, autoantibodies or salivary gland biopsy) in comparison with previous childhood Sjögren’s studies **(Supplementary Table 4)**. However, meeting the criteria may exclude an earlier stage of disease that is not captured through these criteria [28,29]. This is especially significant for Ro-negative children with a high suspicion of having a primary SjS, because it is unclear whether these autoantibodies could be appearing later, although salivary gland biopsy will play a key role in such cases in confirming SjS. In Ro-negative children with a normal histopathological analysis, the disease can be reasonably discarded. But as happens in adults, the criteria are designed to classify and not to diagnose on clinical grounds, and some children could have an abnormal biopsy with focal sialadenitis but not fulfilling the classification criteria (a focus score <1, but > 0) [30]. Only long-term follow-up studies of individuals diagnosed with primary SjS during childhood will confirm whether these patients will develop a full SjS during the follow-up. These studies will be also helpful for defining the prognosis of the childhood-onset SjS. In our cohort, only 1 (0.6%) case showed a lymphoma coincident with the diagnosis of primary SjS. Although lymphoma development has been reported in some children with SjS [31-34], whether the risk is similar to or greater than that in adults is not yet known.

The study has some limitations. With a retrospective design analysing pre-existing data obtained from medical records, a recall bias cannot be discarded. The retrospective use of the ESSDAI score (which was published in 2010) also means that some laboratory parameters were not available at diagnosis in all patients. In addition, the physician assessment and the referral patterns from each centre may influence how systemic disease is scored. The predominant presence of European patients (due to the origin of the project in the EULAR SS Group) could also limit the generalization of the results in non-European populations due to the small size of some ethnic subpopulations, such as Black/African-American patients, and further studies may be necessary to confirm their relevance in these patients.

In summary, childhood-onset primary SjS involves around 1% of patients with primary SjS, with a clinical phenotype dominated by sicca features, parotid enlargement and systemic disease. Our results indicate that no essential difference exists in SS between young-onset and childhood-onset patients, suggesting that the specific features seen in childhood-onset disease may reflect an early stage of SjS. Therefore, childhood-onset SjS should be considered an epidemiological subset with a specific pattern of presentation as happens with other epidemiological subsets (elderly patients, male patients) more than a differentiated disease from adult-onset primary SjS, highlighting the key role of the age at diagnosis on modulating the phenotypic expression of the disease from childhood to elderly ages. Paediatric-specific normative values for these diagnostic tests and paediatric-specific classification criteria are needed, as well as international collaborative studies to better define and understand the natural history of SjS in children, and to determine what features accurately could predict a worse progression.

## Data Availability

Data are available upon reasonable request

## CONFLICT OF INTEREST

None declared.

## ACKNOWLEDGMENTS

Other members of the Sjögren Big Data Consortium that have contributed to this work:

**B. Kostov** (Department of Statistics and Operational Research, Universität Politècnica de Catalunya; Primary Healthcare Transversal Research Group, IDIBAPS, Barcelona, Spain); **I-F. Horvath, A. Szanto** (Division of Clinical Immunology, Faculty of Medicine, University of Debrecen, Debrecen, Hungary); **R. Seror, X. Mariette** (Center for Immunology of Viral Infections and Autoimmune Diseases, Assistance Publique - Hôpitaux de Paris, Hôpitaux Universitaires Paris-Sud, Le Kremlin-Bicêtre, Université Paris Sud, INSERM, Paris, France Paris, France); **Marika Kvarnstrom, M. Wahren-Herlenius** (Department of Medicine, Solna, Division of Experimental Rheumatology, Karolinska Instituted and Karolinska University Hospital, Stockholm); **S. Praprotnik** (Department of Rheumatology, University Medical Centre, Ljubljana, Slovenia); **R. Solans** (Department of Internal Medicine, Hospital Vall d’Hebron, Barcelona, Spain); **G. Nordmark** (Rheumatology, Department of Medical Sciences, Uppsala University, Uppsala, Sweden); **D. Hammenfors, J.G. Brun** (Department of Rheumatology, Haukeland University Hospital, Bergen, Norway); **T. A. Gheita** (Rheumatology Department, Kasr Al Ainy School of Medicine, Cairo University, Egypt); **F. Atzeni** (IRCCS Galeazzi Orthopedic Institute, Milan and Rheumatology Unit, University of Messina, Messina, Italy); **B. Armagan, L. Kilic, U. Kalyoncu** (Department of Internal Medicine, Hacettepe University, Faculty of Medicine, Ankara, Turkey); **T. Nakamura, Y. Takagi** (Department of Radiology and Cancer Biology, Nagasaki University Graduate School of Biomedical Sciences, Nagasaki, Japan); **S. Consani** (Clinica Medica 3- Universidad de la Republica, Hospital Maciel, Montevideo, Uruguay); **F. Olivera Solorzano** (Department of Paediatric Rheumatology, Unidad de Especialidades Médicas. Secretaría de la Defensa Nacional, México DF, México).

## KEY MESSAGES

- Primary SjS is exceptionally diagnosed in children (around 1% of cases).
- Salivary gland enlargement is a prominent feature of childhood-onset primary SjS affecting one out of two children.
- Salivary gland biopsy was positive in 97% of children, and nearly 90% were positive for Ro/La antibodies.
- Childhood primary SjS is undeniably a systemic disease (90% of cases had systemic activity at the time of diagnosis).
- The study highlights the key role of the age at diagnosis on modulating the phenotypic disease expression from childhood to elderly ages.

## Notes

### Competing Interest Statement

The authors have declared no competing interest.

### Funding Statement

The study was supported by Grants Fondo de Investigaciones Sanitarias (MRC, INT15/00085) and ‘Ajut per a la Recerca Josep Font’ (PBZ, Hospital Clinic-Barcelona 2012).

